# Exploring the role of the private sector in tuberculosis detection and management in Lima, Peru: a mixed-methods patient pathway analysis

**DOI:** 10.1101/2023.09.27.23296252

**Authors:** Christoph Wippel, Sheyla Farroñay, Hannah N. Gilbert, Ana Karina Millones, Diana Acosta, Isabel Torres, Judith Jimenez, Leonid Lecca, Courtney M. Yuen

## Abstract

In Latin America, little is known about the involvement of private healthcare providers in TB detection and management. We sought to gain a better understanding of current and potential roles of the private sector in delivering TB services in Peru. We conducted a mixed-methods study in Lima, Peru. The quantitative component comprised a patient pathway analysis assessing the alignment of TB services with patient care-seeking behavior. The qualitative component comprised in-depth interviews with 18 private healthcare providers and 5 key informants. We estimated that 77% of patients initially sought care at a facility with TB diagnostic capacity and 59% at a facility with TB treatment capacity. The lack of TB services at initial care-seeking location was driven by the 41% of patients estimated to seek care first at a private facility. Among private facilities, 43% offered smear microscopy, 13% offered radiography, and none provided TB treatment. Among public sector facilities, 100% offered smear microscopy, 26% offered radiography, and 99% provided TB treatment. Interviews revealed that private providers believed that they offered shorter wait times and a quicker diagnosis, but they struggled with a lack of follow-up systems and communication barriers with the public sector. While expressing willingness to collaborate with public sector programs for diagnosis and referral, private providers had limited interest in treating TB. This study highlights the role of private providers in Peru as an entry point for TB care. Public-private collaboration is necessary to harness the potential of the private sector as an ally for early diagnosis.

## INTRODUCTION

Tuberculosis (TB) has once again surpassed COVID-19 to reclaim its position as the world’s deadliest infectious disease, responsible for approximately 1.6 million deaths in 2021.^1^ Nearly one-third of all people with TB may not receive an accurate diagnosis and suitable care. ^1^ Limited access to healthcare services and insufficient means of detecting cases contribute to delayed or missed diagnoses.^2^ To ensure timely diagnosis for all patients, TB services must be accessible where people with TB seek care.

Patient pathway analyses (PPA) from multiple countries revealed that a sizable portion of people with TB seek care first in the private sector.^3–7^ After this initial care-seeking, many people with TB visit multiple facilities, moving between private and public sectors, before receiving a TB diagnosis.^8–10^ Despite the importance of the private sector in the pathway to diagnosis for many people with TB, many national TB programs engage less with private providers than the public healthcare system. Some PPA studies have revealed a lack of basic information on TB diagnostic service availability in the private sector, as government regulatory agencies do not comprehensively cover private facilities.^3, 7, 11^ Yet in Asia and Africa, engaging the private sector has consistently been shown to increase TB case detection and improve treatment outcomes, and various models for partnership exist.^12^

Despite private healthcare providers playing an increasingly significant role in healthcare delivery in Latin American nations, the literature on the involvement of private healthcare providers in TB detection and management for this region is limited. Studies from Mexico, Nicaragua, and Peru have shown that substantial proportions of people with TB initially seek care in private sector facilities.^13–15^ A study from Brazil found that a relatively small, albeit growing, percentage of TB diagnostic tests were performed in the private sector.^16^ However, among the 78 studies included in a systematic review of public-private partnership models for TB,^12^ only one originated from Latin America, and it focused on the involvement of private pharmacies rather than private health facilities.^17^ We sought to gain a better understanding of current and potential roles of the private sector in TB diagnosis and management in Peru. We therefore carried out a mixed-methods study, combining the first TB PPA in a Latin American country with a qualitative examination of private sector providers’ perspective on their role in TB care.

## MATERIALS AND METHODS

We used a convergent mixed methods design. The quantitative component comprised a PPA.^18^ Qualitative data was collected to elaborate, clarify and explain the quantitative results by exploring the role of private healthcare providers in TB care. While the PPA encompasses all sectors of the healthcare system, the qualitative data focus specifically on the private sector because public sector TB services are in general better understood.^15, 19, 20^ Data were collected between May and October 2022.

### Study setting

Lima, the capital of Peru, is divided into four administrative regions that are each overseen by a regional health authority of the Ministry of Health. This study was conducted in the North Lima region, which comprises nine districts and a population of ∼2.5 million. Peru’s public healthcare sector mainly comprises the Ministry of Health (MINSA) system, which serves people with government insurance provided to those who lack other forms of insurance, and the EsSalud system, which is affiliated with the Ministry of Labor, and which serves people with employer-funded social insurance. The private sector provides services to people who pay out-of-pocket or who possess private insurance. The Ministry of Health establishes national healthcare standards, but it does not actively supervise EsSalud or the private sector.

### Patient Pathway Analysis (PPA)

The study’s quantitative component comprised a PPA, a standardized approach to evaluate the alignment between care-seeking behavior and the TB services.^18^ Data on healthcare-seeking behavior among people with TB was drawn from a previous study from Lima.^15^ Because no information on TB service availability in the private sector was available, we conducted brief surveys with providers working in private healthcare facilities about the availability of radiography, smear microscopy, and TB treatment. For feasibility, we limited the surveys to Carabayllo, one district of North Lima, and extrapolated the results to the other districts. We obtained data on TB service availability in Ministry of Health facilities from the regional health authority (DIRIS Lima Norte) and in the EsSalud network from their public websites. We used World Health Organization TB incidence estimates for 2021^1^ and data on 2021 TB notifications and treatment outcomes for the 2019 and 2020 treatment cohorts from the National TB Program portal;^21^ these data were only available at the national level, so we assumed that they applied to North Lima.

To estimate the number of private sector facilities where a person with TB could seek care, we obtained a list of all registered private healthcare facilities in North Lima from the National Health Regulatory and Supervisory Agency (SUSALUD). We filtered the list to categories that include general medical attention, then excluded those that provided only specialist care (e.g. dental, psychological, ophthalmological). We visited the remaining registered facilities in Carabayllo and determined how many were functioning. We used the ratio of functioning to registered facilities to estimate the total number of functioning private facilities in North Lima.

We categorized Ministry of Health facilities into primary care centers and hospitals. We considered EsSalud and private sector facilities each as a single category because distinctions between different types of facilities were not as standardized. Within each category, we estimated *diagnostic coverage* of smear microscopy and radiography, defined as the proportions of facilities with these capabilities; rapid molecular testing is not widespread in Peru. Combining the published data on initial care seeking location with diagnostic coverage for each facility category, we estimated the percentage of people with TB who had *diagnostic access at initial care seeking*, defined as first seeking care at a facility offering either smear microscopy or radiography. We used a similar approach to estimate *treatment coverage* (the proportion of health facilities that TB treatment) and *treatment access at initial care seeking* (the proportion of people who first sought care at a facility offering TB treatment). We estimated *notification location* (the proportion of people with TB who were diagnosed and notified, by sector) by combining national-level TB notification data with WHO’s 2021 total TB incidence estimates.

We estimated *treatment outcome* (the proportion of people with TB who were successfully treated) by applying treatment success rates for sensitive- and drug-resistant TB to the corresponding notification numbers, and applying this overall treatment success rate to the WHO’s 2021 total TB incidence estimates.

### Qualitative data collection and analysis

We recruited for interviews key informants who were knowledgeable about policies that affect TB services in the private sector and healthcare providers working in the private sector in North Lima. We used existing professional connections of the study team to recruit key informants, which included representatives of the DIRIS Lima Norte, national-level Ministry of Health, SUSALUD, and Socios En Salud, a non-profit non-government organization (NGO) active in the field of TB care. We recruited private sector providers from among those who completed the PPA survey. We purposively sampled an equal number of participants who reported having diagnosed TB within the past year and those who had not. Because our PPA survey sample had a limited number of providers who had diagnosed TB, we employed snowball sampling to reach our initial target for interviews, requesting interviewees to refer other information-rich private health providers practicing in North Lima for participation. In total, we conducted 22 interviews involving 23 participants: 5 key informants, 12 private providers who were a subset of the survey sample, and 6 additional private providers identified through snowball sampling. Eight interview participants were female and 15 were male. All but two of the private providers were medical doctors (one was a nurse, and one was a midwife). Ten survey participants were not interested in being recruited for interviews, and no recruited interview participants declined.

We conducted interviews using semi-structured interview guides (Supplementary File). Interviews were conducted in Spanish by trained Peruvian nurse technicians (SF, DA, IT) working for the NGO Socios En Salud, all of whom had experience in qualitative research. The interviewers did not previously know the participants, except for one key informant who worked for Socios En Salud in a different department. Interviews lasted 30–90 minutes. Three interviews were conducted via videoconference and the rest in person. All interviews were audio recorded, and a note-taker was present during each interview (either one of the other interviewers or CW, a medical doctor).

Interview transcripts were created using artificial-intelligence-assisted software (HappyScribe, Barcelona). Transcripts were checked and corrected by SF and CW. CW translated the transcripts into English; SF provided knowledge about the local context to ensure accurate translation. We employed a content analysis approach to data analysis.^22^ CW open-coded a subset of high-quality transcripts, discussed the results with HNG (a medical anthropologist) and CMY (an epidemiologist), and developed a final codebook with 34 codes. CW coded the entire dataset using Dedoose v9.0.17 (SocioCultural Research Consultants LLC, Los Angeles, CA). CW, HNG, and CMY analyzed the coded data using an inductive approach, generating a preliminary set of descriptive categories, which were refined through iterative examination. LL helped with interpretation.

We integrated quantitative and qualitative results by overlaying the qualitative insights onto the PPA indicators using a joint display technique.^23^ This approach allowed us to understand how the private sector might serve as a facilitator or barrier to TB care at different points along the patient pathway.

### Ethical considerations

This study received approval from the Socios En Salud Institutional Research Ethics Committee and was exempted by the Harvard Medical School Institutional Review Board. We obtained verbal consent from survey respondents and written consent from interview participants.

## RESULTS

Figure 1 shows the quantitative PPA results and qualitative themes structured around the main components of the patient pathway. Tables 1-3 show supporting quotes for qualitative themes corresponding to components I through IV of the patient pathway illustrated in Figure 1.

**Figure 1.**
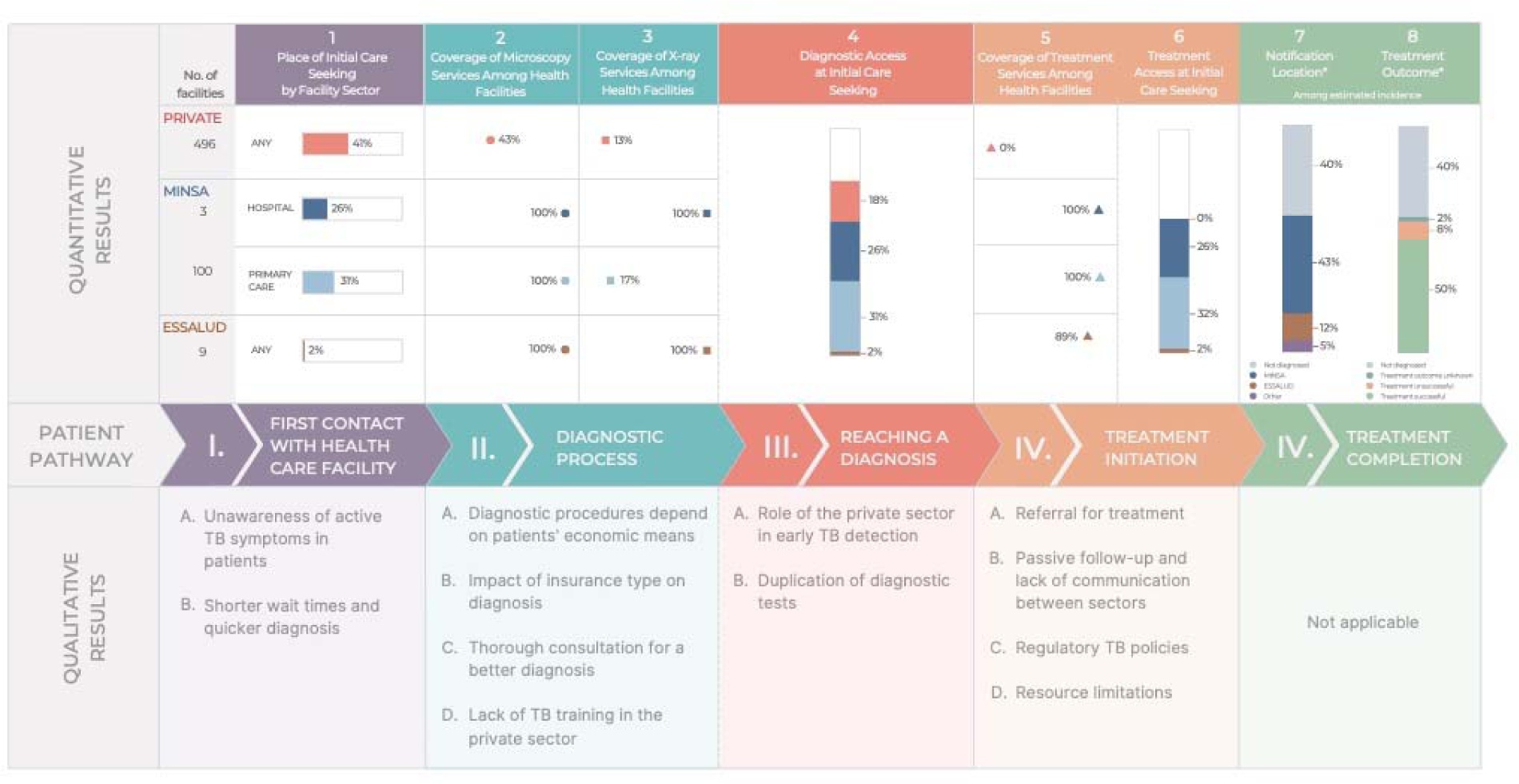
Joint display of integrated results presented longitudinally across the patient pathway. Quantitative results are stratified by sector, considering the private sector, the public system operated by the Ministry of Health (“MINSA”), and the public system operated by the Ministry of Labor (“EsSalud”). Notification data (*) were used from 2021, when case notifications had decreased by approximately 20% during the COVID-19 pandemic; therefore, the percentage of missed people with TB is higher than in prior years.

**Table 1:** Quotes illustrating themes related to first contact with health facilities (step I in patient pathway)

| Theme | Quote |
| --- | --- |
| I-A. Unawareness of active TB symptoms in patients | *“I think that the main reason why a person would come to a private establishment with tuberculosis is to learn about their disease. They might confuse the symptoms and think that it is another respiratory problem and not TB, because most people know that TB is managed in public centers.”*<br>*(Interview 10, Private provider who had diagnosed TB in the past year)* |
| I-B. Shorter wait times and quicker diagnosis | *“If there is suspicion [for TB], we request a sputum BK [smear microscopy] test. This BK test can be done privately or publicly at MINSA. Publicly, it's free, but the results take a month. [...] The patient arrives at the public sector with a diagnosis, and what does the TB program do? They start the treatment. You skipped the whole month or two months that the pulmonologist would have taken to get the x-rays at MINSA, the CT scan, and the BK test. That time was shortened, and you already have the diagnosis and can start treatment.*<br>*(Interview 18, Private provider who had diagnosed TB in the past year)* |

**Table 2:** Quotes illustrating themes related to the diagnostic process and reaching a diagnosis (steps II and III in patient pathway)

| Theme | Quote |
| --- | --- |
| II-A. Diagnostic procedures depend on patients' economic means | *"Normally in all private centers, we have more access to blood, ultrasound, and radiological tests, because the person who goes to a [private] clinic usually has the economic means to afford to afford it. So, we reach a diagnosis more easily because we ask for everything that is needed, they do all the tests, and the diagnosis is made quickly." (Interview 17, Private provider who had diagnosed TB in past year)* |
|  | *"The cost of the consultation is low and they make the most money with the laboratory. For example, with [facility name], I know of many patients I have seen with TB that have gone through [facility name] and they all come with spirometry and many expensive tests as well. And they did not do a smear because it was very cheap." (Interview 1, Key informant from NGO)* |
| II-B. Impact of insurance type on diagnosis | *"If I were in a clinic that had everything, and the patient had an EPS [private insurance] that covers everything, I would order everything, from a sputum BK [smear microscopy] to a tomography scan. But we are talking about the poorest portion of the population, we have to think carefully about what we order for them because sometimes the patient does not have the means to pay for everything. [...] I take a good medical history and a physical exam, and if they have respiratory symptoms of a contagious disease such as tuberculosis, I order them to get a serial sputum BK – usually, because those are the cheapest, the sputum BK and a chest x-ray." (Interview 10, Private provider who had diagnosed TB in past year)* |
| II-C. Thorough consultation for a better diagnosis | *"Ideally, they [healthcare providers] take their time to take a good medical history for the patient. If they achieve this, it's already 80% – the diagnosis is 80% in the medical history. [...] They have to dig for things, and then they will find out what the problem is." (Interview 2, Private provider who had diagnosed TB in past year)* |
| II-D. Lack of TB training in the private sector | *"Here, in this establishment, I have not received any training, neither for diagnosis nor for management of TB. I have been practicing here in the country since [date] 2020. [...] Until now, I have not had any training here, at least not with respect to TB." (Interview 5, Private provider who had not diagnosed TB in past year)* |
| III-A. Role of private sector in early TB detection | *"Well, in our case as a country, as I mentioned, one of the main advantages that could be considered for their case is early detection. That is, the ability to detect or at least identify probable cases for quick referrals. I mean, in that they could contribute a lot." (Interview 22, Key informant from Ministry of Health)* |
| III-B. Duplication of diagnostic tests | *"There may be requests for sputum smear tests to private laboratories, but we do not know if they are truly capable of providing that type of test. [...] It has always been considered that if diagnoses are made in the private sector, they must be validated because if they are not evaluated by the [National Health Institute] that gives a certain guarantee, we will not have the confidence in* |
|  | *knowing that those diagnoses are truly what they are.” (Interview 22, Key informant from Ministry of Health)* |

**Table 3:** Quotes illustrating themes related to treatment initiation (step IV in patient pathway)

| Theme | Quote |
| --- | --- |
| IV-A. Referral for treatment | *“Unfortunately, the private sector refers patients to the Ministry of Health system for treatment because they don't have the treatment regimens. It would be ideal to give the patient a week of treatment and instruct them to follow up at a nearby health center for monitoring, but that's not something that we are currently doing.” (Interview 20, Private provider who had diagnosed TB in the past year)* |
| IV-B. Passive follow-up and lack of communication between sectors | *“Patients usually reach out to me... and update me on the progress of the tests and results or if they have been cleared. So it is the opposite – patients reach out to me rather than me reaching out to them.” (Interview 13, Private provider who had not diagnosed TB in the past year)* |
| IV-C. Regulatory TB policies | *“I am not familiar with any regulations in the private sector for TB. What I do know is that all TB cases should be referred to the nearest Ministry of Health facility.” (Interview 18, Private provider who had diagnosed TB in the past year)* |
|  | *“There are private facilities that, depending on the level of complexity, are definitely capable of treating tuberculosis [...] They should treat tuberculosis, they are prepared, they even have their pulmonologist, and they have their areas. That is why this should be only reserved for the [more complex] clinics, because now they have to comply with new regulations.” (Interview 3, Private provider who had not diagnosed TB in the past year)* |
| IV-D. Resource limitations | *“And seeing it from the point of a private provider, I think it would be a bit difficult, financially speaking [to have a pulmonologist on standby in case we need one]. So, I am seeing it from the perspective of how much that would cost me.” (Interview 11, Private provider who had not diagnosed TB in the past year)* |

### I. First contact with healthcare facility

In North Lima in 2022, there were 1,687 registered private health facilities, of which we estimated that 859 provided general medical services. In the district of Carabayllo, we were able to find 26 functioning private facilities, compared to the 45 registered. Applying a similar ratio throughout North Lima yielded an estimated 496 functioning private facilities. Public health facilities in North Lima with general medical services included 100 Ministry of Health primary care centers, 3 Ministry of Health hospitals, and 9 EsSalud facilities. A previous study from Lima found that 41% of people with TB first sought care in the private sector, 31% at Ministry of Health primary care centers, 26% at Ministry of Health hospitals, and 2% at EsSalud facilities.^15^

#### A. Lack of awareness of TB symptoms

Private healthcare providers perceived that although the general population is aware that treatment for TB is provided in public programs, people may not recognize their symptoms as indicative of TB and therefore initially seek care in a private facility, which may be the nearest or most convenient facility.

#### B. Shorter wait times and quicker diagnosis

Private healthcare providers viewed shorter wait times for appointments and during healthcare visits, as well as being able to obtain diagnostic results faster as both reasons why patients seek care in the private sector and the strength of their services. By providing patients with earlier appointments, quicker test results, and more thorough consultations, private providers believed that they could make a TB diagnosis sooner and thereby reduce delays to treatment initiation compared to the public sector.

### II. Diagnostic process

Providers from 23 of the 26 (88%) private facilities in Carabayllo agreed to take the survey on TB service availability. Smear microscopy was offered in 43% of the surveyed private facilities and 100% of Ministry of Health primary care centers, hospitals, and EsSalud facilities. Radiography was offered at 13% of private facilities, 17% of Ministry of Health primary care centers, and 100% of Ministry of Health hospitals and EsSalud facilities.

#### A. Diagnostic procedures depend on patients’ economic means

Private providers believed that one of the strengths of the private sector is the ability to arrive at a faster diagnosis in part because of the availability of more advanced technologies. Collaborations with private laboratories and more advanced diagnostic technologies such as MRI, CT scans, and GeneXpert in better-equipped facilities can promote faster diagnosis. However, the accessibility of these services is dependent on the patient’s financial means, as they typically pay out-of-pocket. A key informant from an NGO mentioned that certain private facilities may bypass basic tests like smear microscopy and immediately proceed with more advanced, and sometimes unnecessary, tests.

#### B. Impact of insurance type on diagnosis

While some private providers indicated that patients’ insurance status did not affect the quality or type of services they received since services were all paid for out-of-pocket, others noted significant disparities in diagnostic services depending on insurance coverage and the patient’s economic means. These providers explained that they tended to offer their patients with private insurance more advanced and expensive tests, while those without insurance or with state insurance were more likely to be offered basic and cheaper tests.

#### C. Thorough consultation for a better diagnosis

The majority of private providers interviewed exhibited familiarity with the clinical indicators that raise suspicion of active TB. They noted that combining these with a thorough social history – including details such as socioeconomic status, living conditions, and social contacts – considerably aids the diagnostic process.

#### D. Lack of TB training in the private sector

However, private healthcare facilities do not provide any training for their staff on TB, and there are no external training programs available for private providers. As a result, private providers’ knowledge of TB is largely based on their prior medical training in academic institutions or their experience working in the public sector. Some private providers reported engaging in self-training out of personal interest.

### III. Reaching a diagnosis

Overall, we estimate that 77% of people with TB initially sought care at a facility with TB diagnostic capacity (sputum smear microscopy or radiography). The lack of TB services at the initial care-seeking location was driven by the people who initially went to a private facility, since all public health facilities offered at least smear microscopy. Of the surveyed private providers, 35% reported that they had diagnosed at least one person with TB in the past year.

#### A. Role of private sector in early TB detection

Private providers agreed that the private sector’s main role lies in the early detection and diagnosis of TB as opposed to treatment. They perceived the availability of resources, speed of diagnosis, and access to cutting-edge technologies as strengths of the private sector. A key informant from the Ministry of Health agreed that there was value in the private sector’s contribution to early TB detection and expressed a general openness to expand the role of the private sector in this capacity. Some providers proposed expanding the private sector’s early detection potential with more widespread use of advanced diagnostic tests, such as GeneXpert, or by implementing a screening program that would test every patient with respiratory symptoms for TB.

#### B. Duplication of diagnostic tests

Some private providers described having diagnosed patients with TB and referred them to the public sector, only to have the diagnostic test results rejected by the public sector and repeated, causing distress to the patient. The Ministry of Health’s key informant explained that this stems from a lack of oversight and validation of private laboratories, which are not supervised or monitored as part of the public network, leading to lower levels of confidence in their results.

### IV. Treatment initiation

None of the private providers offered treatment for TB, while treatment was available at all Ministry of Health facilities and 89% of the EsSalud facilities. Thus, we estimate that 59% of people with TB had access to treatment services at their initial care location.

### A. Referral for treatment

There was a clear consensus that the private sector does not provide TB treatment, with interviewees citing legal prohibitions, medication inaccessibility, and the belief that the public program is performing well. While many private providers asserted that the private sector’s responsibility ends with referral of people with TB to the public sector, some suggested that they should have the option of providing initial treatment after diagnosis until the patient reaches the public sector to avoid delay in starting treatment.

#### B. Passive follow-up and lack of communication between sectors

Upon referral to the public sector, the responsibility for treatment often shifts to patients themselves. For most private providers, the referral process is viewed as a handover to the public program, with the expectation that the patient will receive appropriate treatment. There are no standardized systems for sharing patient information from the private to the public sector after a patient is diagnosed with TB and referred, or to inform the referring private facility that a patient has arrived and initiated treatment in the public facility. Private providers expressed the desire for better feedback from the Ministry of Health system about the status of referred patients. A digital platform for sharing patient information exists for public facilities, but private facilities are currently excluded. During the COVID-19 pandemic, this system was made accessible to private facilities for the purposes of sharing vaccine-related data with the public sector, which raised the question of why something similar could not be achieved for TB.

#### C. Regulatory TB policies

Private healthcare providers believed that regulatory policies prohibit the private sector from treating TB patients, although they did not generally know the details of the policies.

However, the NGO key informant mentioned that regulatory policies provide for a mechanism that could allow private facilities to enter into an agreement with the Ministry of Health to treat drug-sensitive TB. However, the private facility would have to fulfill strict requirements at their own cost, including specialized staff, provisions for contact tracing, follow-ups, supplemental services, and biosafety. No private facility had entered into such an agreement at the time of this study. High levels of bureaucracy and few financial incentives were mentioned as possible reasons for the low levels of interest in this mechanism.

#### D. Resource limitations

Many private providers felt it would be difficult for their facilities to deliver quality TB care, given the diverse services required to address patients’ needs. Specialist staff like pulmonologists, nutritionists, and psychologists are often not part of the regular staff of private facilities, and it can be financially challenging to retain them on standby. In addition, a lack of adequate space and biosafety infrastructure were mentioned as possible limitations for the provision of treatment in the private sector.

### V. Treatment completion and outcome

At the national level, the WHO estimated 44,000 cases in 2021. A total of 26,437 cases were reported (60% of estimated incidence): 71% by Ministry of Health facilities, 20% by EsSalud facilities, and 9% by other government sectors, such as penitentiaries, the military, and the police ^21^. No cases were reported by the private sector. Among people who initiated treatment, 84% were treated successfully. Thus, an estimated 50% of all people with TB were treated successfully given that 40% were not diagnosed.

## DISCUSSION

Our study highlights the role of the private sector in the diagnosis of TB in North Lima, Peru, and the need for enhanced public-private collaboration. Private facilities greatly outnumber public facilities in this setting, and many people with TB use the private sector despite having to pay out-of-pocket.^15^ However, the limited availability of TB diagnostic services in the private sector meant that around a quarter of people with TB are estimated to have initially sought care at locations without TB diagnostic services. While the private providers we interviewed believed that they could offer shorter wait times and a quicker diagnosis, they placed the responsibility for treatment on the public sector, citing human resource and space limitations, as well as current regulatory policies. However, private providers struggled with a lack of standardized systems for following up people referred into the public sector.

Our findings highlight the potential benefits of closer collaboration between private providers and the Peruvian national TB program to improve TB detection. While studies from other settings have generally documented willingness of private providers to engage with the national TB program,^24^ some studies have documented low motivation and even resistance to transferring patients to public sector facilities.^25, 26^ Providers in our study were comfortable with referring TB patients to the public sector for treatment and did not make a clear call for treatment in the private sector. Thus, public-private partnership efforts in Peru should focus on strategies to improve TB diagnosis in the private sector and facilitate efficient referral to the public sector.

While our qualitative findings depicted a generally positive outlook concerning TB diagnosis in the private sector with private providers believing they were performing well, our quantitative results indicated that most private facilities lacked TB diagnostic capabilities, and a minority of surveyed private providers had diagnosed a case of TB in the past year. In part, this inconsistency is likely attributable to the characteristics of our interview sample: providers who agreed to interviews were predominantly from facilities with diagnostic capacities, and we purposively sampled providers such that half had diagnosed TB recently. Even so, the interviewed providers who had not recently diagnosed TB did not highlight challenges or limitations to TB diagnosis in the private sector. This suggests either that they were unaware of these limitations, or that social desirability bias prevented them from discussing these issues. Studies of the quality of private sector TB diagnostic services in Asian and African countries have found wide variation in quality within settings, and have documented low TB testing rates even for people with typical TB symptoms.^27^ It is likely that such variation in quality exists in Peru and other Latin American countries as well, and that diagnoses are being missed in the private sector.

The main barrier that hinders effective cooperation between the private and public sectors in Peru is the lack of communication and information sharing. Effective strategies for public-private partnership include the national TB program providing technical support to private providers and establishing formal collaborative relationships with defined expectations.^12^ In Peru, the Ministry of Health could help harness the potential of the private sector by providing training for private providers and formally recognizing private facilities that offer high-quality diagnostic services so that public facilities can accept their diagnostic test results without repeating them. Successful public-private partnerships have also highlighted the importance of formal feedback mechanisms that inform private providers about what happens to their patients who have been referred to the public sector for treatment.^28, 29^ In Peru, a simple way to formalize the referral process would be to give private providers a standardized paper referral form that would be recognized by public facilities. Granting them access to the electronic system through which public facilities to refer and monitor patients would enable both referral and feedback between sectors.

One strength of our study is the use of in-depth interviews with private healthcare providers in combination with a PPA. This approach provides insight into what happens when people with TB seek care in the private sector, which was poorly understood in our setting, as in other Latin American countries. There are several limitations to our research. Firstly, because the list of private facilities was not updated to reflect closures and data were not available on TB services in the private sector, we collected data by directly identifying facilities and conducting surveys of TB service availability. This was only feasible to do for a single district. Carabayllo is one of the larger but less urbanized districts of North Lima, so extrapolating the findings from this district to others may underestimate service availability in the private sector in North Lima. Additionally, we relied on healthcare-seeking data from a prior study of people with TB in Lima,^15^ which included only people being treated within the Ministry of Health system. People treated in the EsSalud system are more likely to have utilized EsSalud facilities as well as private facilities due to existing insurance partnerships. Given that EsSalud facilities have TB services and private facilities may or may not, it is unclear how including these individuals would change the results of the PPA, although the magnitude of any changes would likely be small since only 20% of people with TB are treated in the EsSalud system.^21^

In conclusion, our study highlights the role private providers play as an entry point of care for a sizable portion of people with TB, even in a country where treatment is restricted to the public sector. Improving linkages between the private and public sectors by establishing a formal referral system and improving communication between sectors could lead to higher case detection rates and improved continuity and timeliness of care for people with TB. To harness the potential of the private sector in TB diagnosis in Peru, it is necessary to begin a dialogue between the public and private sectors in order to explore possible public-private partnership models.

## Authors

Christoph Wippel, Harvard Medical School, Boston, USA, Sheyla Farroñay, Socios En Salud Sucursal Perú, Lima, Peru

Hannah N. Gilbert, Harvard Medical School, Boston, USA, Ana Karina Millones, Socios En Salud Sucursal Perú, Lima, Peru, Diana Acosta, Socios En Salud Sucursal Perú, Lima, Peru

Isabel Torres, Socios En Salud Sucursal Perú, Lima, Peru, Judith Jimenez, Socios En Salud Sucursal Perú, Lima, Peru, Leonid Lecca, Socios En Salud Sucursal Perú, Lima, Peru, Courtney M. Yuen, Brigham and Women’s Hospital, Boston, USA

## Data Availability

Data are available upon reasonable request to the authors

