## Supplementary File for "Exploring the role of the private sector in tuberculosis detection and management in Lima, Peru: a mixed-methods patient pathway analysis"

Interview guide #1 – Key informants

**Research project: Tuberculosis diagnosis and treatment in the private sector in Lima, Peru**

Specific aims:

-Understanding the role of the private health care sector in the treatment of TB.

-Identifying policies for TB treatment concerning the private sector

-Finding out whether there are barriers for the private sector in the provision of diagnostic and treatment services for TB.

Intended interviewees:

Key informants who are knowledgeable with the national TB treatment policies

Introduction:

*Thank you for doing this interview with me today. This interview is part of a research project looking at the treatment pathway of patients with tuberculosis (TB) with a particular focus on patients who initially seek care in private sector facilities. The interview will take approximately 60-90 minutes, depending on how much we have to talk about. Everything we talk about will be treated confidentially and you are free to decline to answer any of the questions at any time. The interview will be audio-taped, and I will also take some notes during our conversation. Do you have any questions before we start?*

*1. Please tell me about your professional role as it relates to TB?*

*2. In the context of your TB work, what kind of interactions have you had with private health care providers? Probe for examples.*

*(Only ask this question if interviewee is not a health care provider)*

*3. What are some barriers to professional communication between providers in the public and private sectors regarding TB services?*

*4. What would encourage more communication between those public and private providers?*

*5. From your view, what are the responsibilities that a private provider has if a patient comes to them, and they suspect this person is living with TB?*

*Probe for detailed procedure until patient is treated.*

*Follow up: Please share any examples of this process that you have encountered.*

*Follow up: What financial considerations do private providers have to consider when diagnosing and treating people with TB?*

*Probe:* What are some possible incentives? Any disincentives? Probe for examples.

*6. What are the provisions in the national health policy for TB diagnosis and treatment in the private health care sector?
Follow up: What are some advantages to having those provisions in place?
Follow up: What are some disadvantages to those provisions related to the private sector?*

*7. Please tell me what you understand about how the process works for private health care providers to obtain TB medications. Probe for examples.*

*8.. How has this worked in your view?*

*Probe for effects on patients.*

*9. What role do you believe the private sector should play in the diagnosis of TB?*

*Follow up: What role should they play in treatment?*

*Follow up: In your view, what should be done?*

*10. In your view, what role, if any, has COVID-19 played in the practices and procedures of TB care in the private sector?*

*11. Is there anything else that I should know to better understand how the private sector works here in terms of TB diagnosis and treatment?*

*12. Do you have any additional thoughts to add about how you would like to see the private sector operate in the TB sphere?*

Interview guide #2 – Private health care providers that treat TB

**Research project: Tuberculosis Patient Pathway Analysis in Northern Lima, Peru, with particular focus on the private health care sector**

Specific aims:

-Understanding the patient care pathways of TB patients who initially seek care at private health care facilities.

-Identifying the availability of TB diagnostic and treatment services at private health care facilities.

-Finding out whether there are incentives/disincentives for the diagnosis of TB at private health care facilities.

Intended interviewees:

Health care workers or administrators of private health care facilities that treat TB.

Introduction:

*Thank you for doing this interview with me today. This interview is part of a research project looking at the treatment pathway of patients with tuberculosis (TB) with a particular focus on patients who initially seek care in private sector facilities. The interview will take approximately 60-90 minutes, depending on how much we have to talk about. Everything we talk about will be treated confidentially and you are free to decline to answer any of the questions at any time. The interview will be audio-taped, and I will also take some notes during our conversation. Do you have any questions before we start?*

Main questions:

*1. How long have you worked in this health care facility?*

*2. What is your role within this health care facility?*

*3. Why do you think people with TB come to your facility to seek care for their illness?*

*4. I would like to understand what happens to patients who come to your facility with respiratory symptoms. What usually happens first? And then?*

*Probe until process is complete.*

*If participant has not yet answered the following, probe further:*

*I have a few more questions about what happens when someone presents with respiratory symptoms.*

*How do you gather the patient’s social history? What kinds of questions do you ask these patients? How is that information helpful to you?*

*What kind of medicine do you provide to these patients when they leave the clinic?*

*5. How do you diagnose TB at your health care facility?*

*Probe about X-ray, sputum testing and drug resistance testing.*

*6. What happens after you diagnose someone with TB at your facility?*

*Follow up: Do you report cases to the national TB program? How do you go about doing that?*

*If they treat patient there:*

*6a. Where do you receive the medication that you use treat your patients?*

*6c. In your view, what does your clinic do really well for TB patients in your care?*

*6d. What are some limitations that you experience?
Follow up: What would help you overcome those limitations?*

*7. Is there any situation where you would refer the patient rather than continue treatment at your facility?*

*If they refer at any point:*

*7a. Where do you refer patients? Why there?*

*7b. What is the process for referral? What is that process like for you/the clinic?*

*7c. How do you make sure the patient gets to the correct facility?*

*8. What are the patient’s out-of-pocket expenses during the whole diagnostic process?*

*Follow up: What financial considerations do you have to make as a clinic when someone with TB seeks care at your facility?*

*Follow up: Can you please describe the clinic’s fee structure in caring for these patients?*

*Follow up: What are some financial advantages for the clinic to providing care to someone with TB? What are some financial disadvantages?*

*9. What role do you believe private facilities like yours currently play in the diagnosis of TB?*

*10. What role would you like to see the private sector play in diagnosis? Why? What would help achieve this?*

*11. Please describe the role you think the private facilities like yours plays in the treatment of TB.*

*12. In your view, what role, if any, has the COVID-19 pandemic played in how your clinic approaches the diagnosis and care of patients living with TB?*

*13. In your view, what role should the private sector play in treatment? What are some current barriers? How can those be addressed?*

*14. Is there anything that you think I should know about TB care at your facility?*

*15. Do you have any final thoughts to share about improving TB care in the private sector?*

Interview guide #3 – Private health care providers that do not treat TB

**Research project: Tuberculosis Patient Pathway Analysis in Northern Lima, Peru, with particular focus on the private health care sector**

Specific aims:

-Understanding the patient care pathways of TB patients who initially seek care at private health care facilities.

-Identifying the availability of TB diagnostic and treatment services at private health care facilities.

-Finding out whether there are incentives/disincentives for the diagnosis of TB at private health care facilities.

Intended interviewees:

Health care workers or administrators of private health care facilities that do not treat TB.

Introduction:

*Thank you for doing this interview with me today. This interview is part of a research project looking at the treatment pathway of patients with tuberculosis (TB) with a particular focus on patients who initially seek care in private sector facilities. The interview will take approximately 60-90 minutes, depending on how much we have to talk about. Everything we talk about will be treated confidentially and you are free to decline to answer any of the questions at any time. The interview will be audio-taped, and I will also take some notes during our conversation. Do you have any questions before we start?*

*1. How long have you worked in this health care facility?*

*2. How would you describe your role within this health care facility?*

*3. In your view, what kind of care do you feel your clinic does a very good job providing?*

*4. You said in your survey that you haven’t diagnosed any patients with TB in your facility in the past year. Why do you think that is?*

*5. What happens to patients who come to your facility with respiratory symptoms?*

*Probe: Please walk me through each step. Let’s start with their initial consultation.*

*Probe: How do you gather the patient’s social history? What kinds of questions do you ask these patients?*

*6. What steps do you take if you suspect someone has TB?*

*Probe: Please think about the most recent time that you suspected a TB infection. Can you walk me through each step that you took with that patient?*

*Follow up: What financial considerations do you have to make as a clinic when someone you suspect has TB seeks care at your facility?*

*If they refer at any point:*

*6a. Where do you refer patients? Why do you refer them there?*

*6b. What is the process for referral? What do you think about that process?*

*6c. How do you make sure the patient gets to the correct facility?*

*7. What kind of training/preparation do clinicians at your facility have in recognizing and diagnosing TB? How about in the treatment of TB?*

*8. What role do you believe private facilities like yours currently play in the diagnosis of TB?*

*9. What role would you like to see the private sector play in the diagnosis of TB? Why? What would help achieve this?*

*10. Please describe the role you think the private facilities like yours plays in the treatment of TB.*

*11. In your view, what role should the private sector play in treatment? What are some current barriers? How can those be addressed?*

*12. Is there anything, you want to tell me about improving TB care in the private sector?*
